# Comparative Analysis of GPT-4Vision, GPT-4 and Open Source LLMs in Clinical Diagnostic Accuracy: A Benchmark Against Human Expertise

**DOI:** 10.1101/2023.11.03.23297957

**Authors:** Tianyu Han, Lisa C Adams, Keno Bressem, Felix Busch, Luisa Huck, Sven Nebelung, Daniel Truhn

## Abstract

**Importance:** Medicine is poised for transformation with artificial general intelligence becoming integral to almost all clinical environments. Currently, the performance of multimodal AI, specifically one powered by GPT-4, in real clinical cases remains uncharted.

**Objective:** To ascertain whether GPT-4V can consistently comprehend complex diagnostic scenarios through both imagery and textual data.

**Design:** A selection of 140 clinical cases from the JAMA Clinical Challenge and 348 from the NEJM Image Challenge were used. Each case, comprising a clinical image and corresponding question, was processed by GPT-4V, and responses were documented. The significance of imaging information was assessed by comparing GPT-4V’s performance with that of four other leading-edge large language models (LLMs).

**Main Outcomes and Measures:** The accuracy of responses was gauged by juxtaposing the model’s answers with the established ground truths of the challenges. The confidence interval for the model’s performance was calculated using bootstrapping methods. Additionally, human performance on the NEJM Image Challenge was chronicled, reflected by the choice percentage selected by challenge participants.

**Results:** GPT-4V demonstrated superior accuracy in analyses of both sources, achieving 73.3% for JAMA and 88.7% for NEJM, notably outperforming text-only LLMs such as GPT-4, GPT-3.5, Llama2, and Med-42. Remarkably, both GPT-4V and GPT-4 exceeded average human participants’ performance at all complexity levels within the NEJM Image Challenge.

**Conclusions and Relevance:** GPT-4V has exhibited considerable promise in clinical diagnostic tasks, surpassing the capabilities of its predecessors as well as those of human experts. However, while its proficiency in identification tasks is commendable, it requires further refinement in decision-making and strategic planning. Despite these encouraging results, such models should be adopted with prudence in clinical settings, serving to augment rather than replace human discretion. Continual research is imperative to fully evaluate the potential impact on patient care.

## Introduction

In the ever-evolving field of healthcare, the integration of artificial intelligence with clinical methods has the potential to reshape clinical practice. The transition from specialized to multimodal models, such as OpenAI’s recently launched GPT-4V(ision), signals a transformative phase. The fusion of linguistic and visual capabilities may advance usability of such models in clinical routine^1^.

The question that needs to be addressed is: “How effective are large language models (LLMs), in a real-world clinical setting?”.^2^ To answer this question, this article provides a quantitative assessment of GPT-4V’s capabilities in the field of multimodal medical diagnostics. Using clinical cases from JAMA and the New England Journal of Medicine Clinical Challenges, diagnostic accuracy of GPT-4V is assessed and compared with human expertise, with its predecessors, and with state-of-the-art open-source models. These clinical challenges, created for healthcare professionals, assess their capacity to comprehend medical situations, combine evidence and deduce suitable conclusions over a wide area of medical expertise. This approach offers a more sophisticated measurement of clinical reasoning compared to conventional assessment tools like USMLE questions that are geared towards medical students.

## Methods

For clinical case descriptions starting from 2017, we extracted questions, images, and answer choices from JAMA^1^ (n=140) and NEJM^2^ (n=348), see Figure 1 a and b. For the NEJM questions we additionally extracted statistics for answers given by human users of the website. Case descriptions along with provided answer choices were fed into the models GPT-4, GPT-3.5 (both by OpenAI), Llama2 (by Meta), and into Med-42, a model fine-tuned for medical use based on Llama2. The models were asked to provide the correct answer based on the case description and the answer choices. For GPT-4V we provided the images alongside the case description.

**Figure 1:**
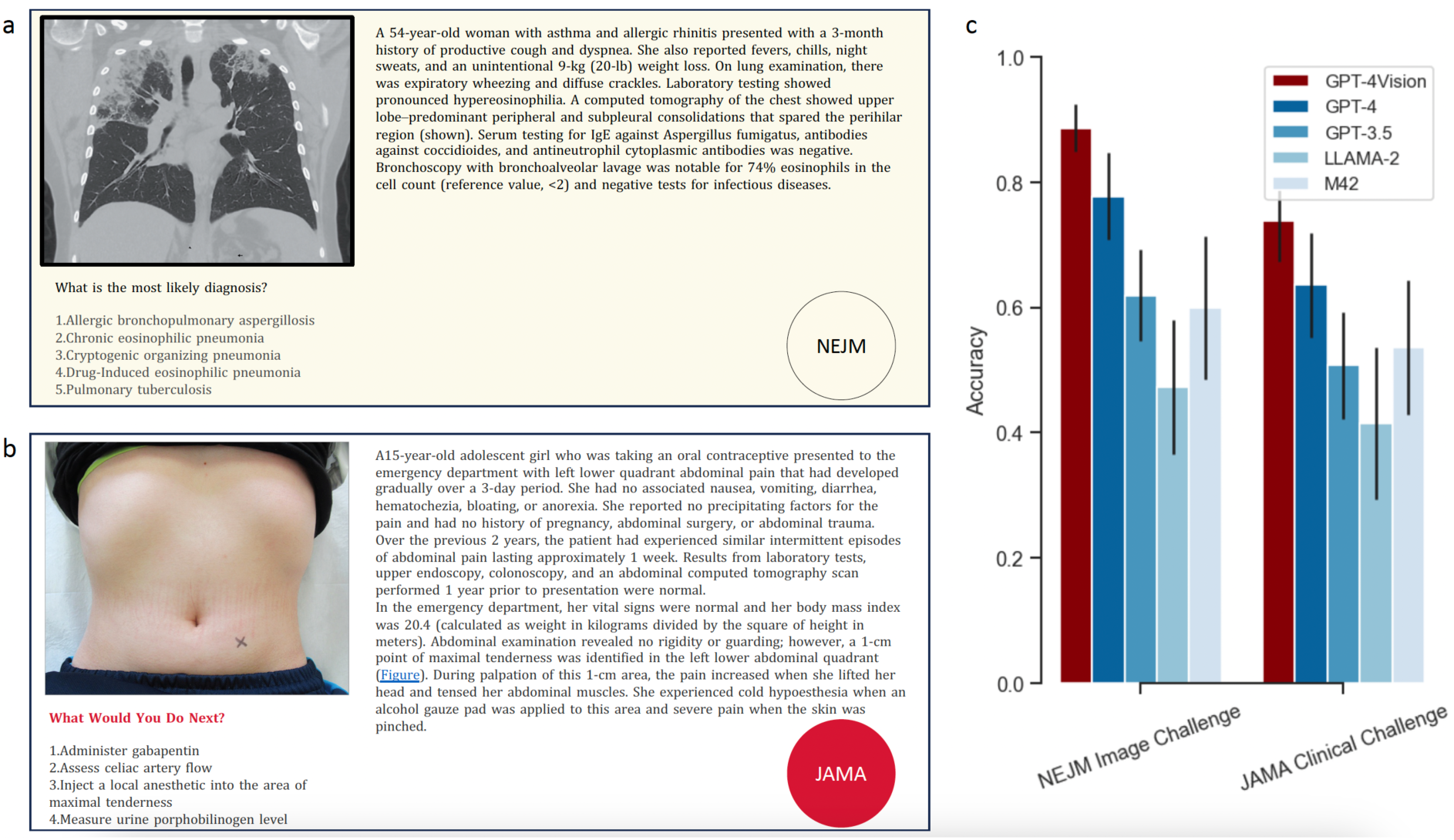
Illustrative examples of the clinical case descriptions from JAMA (a) and NEJM (b) and accuracy of the prorietary models GPT-4 Vision, GPT-4, GPT-3.5, and of the open-source models Llama2 and Med24 in answering the questions (c).

## Results

GPT-4V consistently achieved the highest accuracy, followed by GPT-4 (73.3% vs. 63.6% for JAMA and 88.7% vs. 77.8% for NEJM), see Figure 1c. GPT-3.5 and the open-source model Med-42 performed similarly (50.7% vs. 53.6% for JAMA and 61.7% vs. 59.9% for NEJM). Llama2 exhibited the lowest performance among the tested models (41.4% for JAMA and 47.1% for NEJM). When stratified along question difficulty as measured by the percentage of correct answers provided by human readers of NEJM, these results were confirmed across all difficulty levels, see Figure 2. Notably, GPT-4V and GPT-4 outperformed human readers across all difficulty levels.

**Figure 2:**
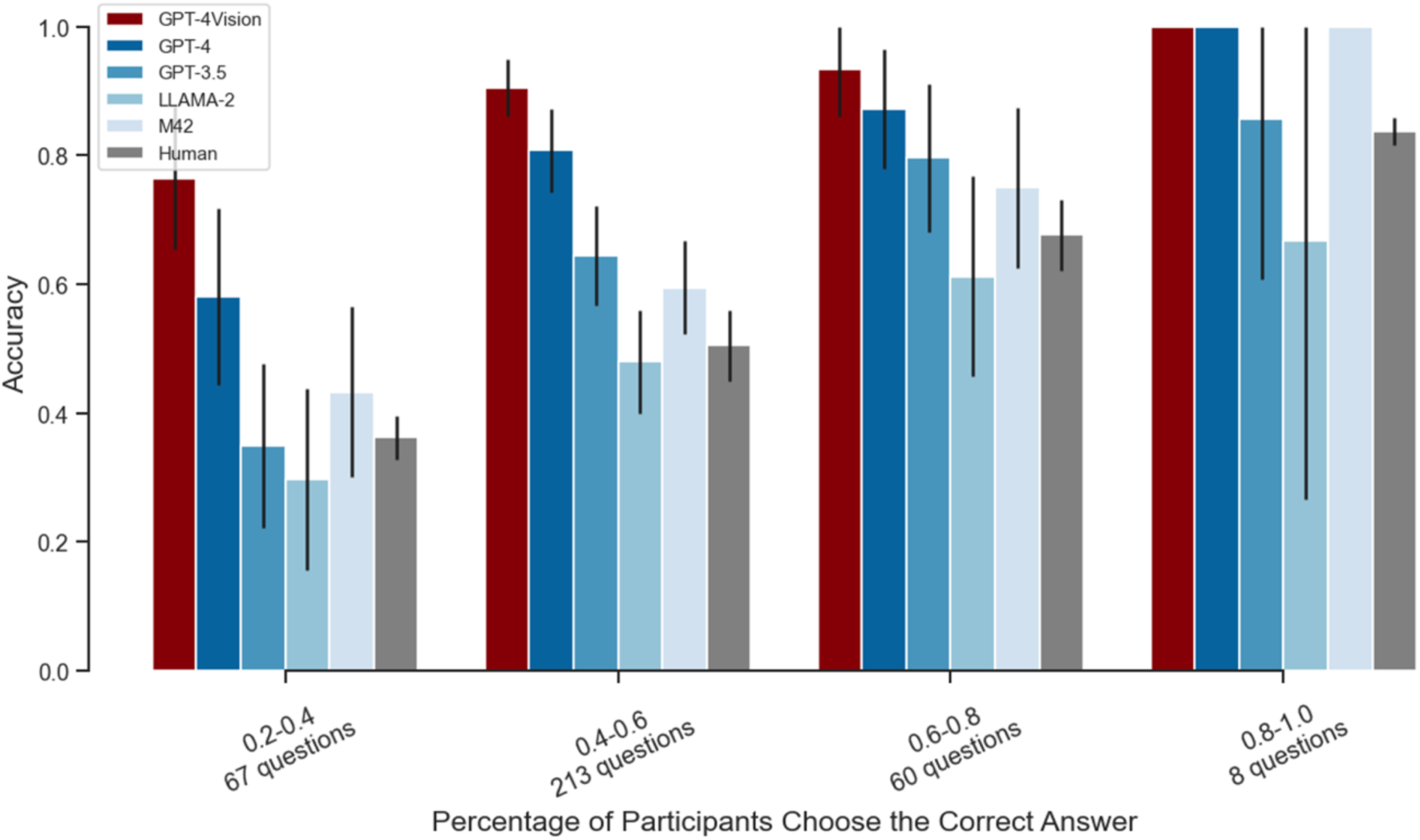
For the NEJM clinical case descriptions, statistics about the accuracy of human readers were provided. We stratified the question difficulty in four categories and evaluated the performance of the LLMs within these categories.

## Discussion

In both the JAMA and NEJM ^3^ Clinical Challenges, GPT-4V demonstrated a significant improvement of more than 10% in performance compared to its predecessors GPT-4 and GPT-3.5, as well as open-source models LLAMA-2 and M42. It also surpassed the accuracy of human diagnosis. Although GPT-4V demonstrated superior diagnostic capabilities, especially in NEJM’s “What is the condition?” format, it encountered relative challenges with JAMA’s forward-thinking query, “What would you do next?”. This discrepancy indicates that while GPT-4V is skilled in identification tasks, further refinement is necessary for its decision-making abilities and planning, a known limitation of current LLMs.^3^ The adaptability and universality of these models across various domains is highlighted by the fact that GPT models did not go through initial training specifically for the medical domain. Although the findings are promising, caution should be exercised as diagnostic accuracy is just one aspect of clinical practice. The integration of AI models must consider their roles in varied clinical scenarios and the broader ethical implications. In summary, although GPT-4V shows promising results in structured clinical tasks, more research is needed to evaluate its overall impact on patient care, and if clinicians use GPT, it should be as a complementary tool, not a replacement for human judgment.^4^

## Data Availability

The data that support the findings of this study are openly available (https://jamanetwork.com/collections/44038/clinical-challenge and https://www.nejm.org/case-challenges). Specific data related to AI model responses can be accessed freely, ensuring transparency and reproducibility of the research.

## Footnotes

1 https://jamanetwork.com/collections/44038/clinical-challenge

2 https://www.nejm.org/case-challenges

